# Development and validation of the RCOS prognostic index: a bedside multivariable logistic regression model to predict hypoxaemia or death in patients with SARS-CoV-2 infection

**DOI:** 10.1101/2021.03.29.21254393

**Authors:** Gerardo Alvarez-Uria, Sumanth Gandra, Venkata R Gurram, Raghu P Reddy, Manoranjan Midde, Praveen Kumar, Ketty E Arce

## Abstract

Previous COVID-19 prognostic models have been developed in hospital settings, and are not applicable to COVID-19 cases in the general population. There is an urgent need for prognostic scores aimed to identify patients at high risk of complications at the time of COVID-19 diagnosis. The RDT COVID-19 Observational Study (RCOS) collected clinical data from patients with COVID-19 admitted regardless of the severity of their symptoms in a general hospital in India. We aimed to develop and validate a simple bedside prognostic score to predict the risk of hypoxaemia or death.

4035 patients were included in the development cohort and 2046 in the validation cohort. The primary outcome occurred in 961 (23.8%) and 548 (26.8%) patients in the development and validation cohorts, respectively. The final model included 12 variables: age, systolic blood pressure, heart rate, respiratory rate, aspartate transaminase, lactate dehydrogenase, urea, C-reactive protein, sodium, lymphocyte count, neutrophil count and neutrophil/lymphocyte ratio. In the validation cohort, the area under the receiver operating characteristic curve (AUROCC) was 0.907 (95% CI, 0.892-0.922) and the Brier Score was 0.098. The decision curve analysis showed good clinical utility in hypothetical scenarios where admission of patients was decided according to the prognostic index. When the prognostic index was used to predict mortality in the validation cohort, the AUROCC was 0.947 (95% CI, 0.925-0.97) and the Brier score was 0.0188.

If our results are validated in other settings, the RCOS prognostic index could help improve the decision making in the current COVID-19 pandemic, especially in resource limited-settings.

## Introduction

Infection with severe acute respiratory syndrome coronavirus 2 (SARS-CoV-2) has high morbidity and mortality [1]. Since late 2019, the rapid spread of SARS-CoV-2 has put an enormous pressure on national health systems worldwide.

The clinical spectrum of coronavirus disease 2019 (COVID-19) produced by SARS-CoV-2 is wide. In a large study from China including 72314 cases, 81% had a mild disease, 19% had a severe disease with deterioration of the respiratory function, and 2.3% died [2]. In many medical domains, prognostic multivariable prediction models have been developed with the aim to help healthcare professionals in their decision making [3]. To date, more than 100 COVID-19 prognostic models have been reported [4]. However, the vast majority of them have been developed in overwhelmed hospital settings from developed countries where the mortality and the proportion of patients with severe disease was high, and they might not be applicable to COVID-19 cases in the general population.

The objective of this study was to develop and validate a pragmatic prognostic score to predict the risk of mortality or hypoxaemia in patients with COVID-19 who were admitted in a hospital regardless the severity of their symptoms. We hypothesized that the population of the study is similar to the general population of COVID 19 cases, and that the prognostic score could be applied in non-hospital settings, where the majority of cases are mild and the proportion of severe cases is relatively small.

## Methods

### Source of data and participants

The Rural Development Trust (RDT) COVID-19 observational study (RCOS) is a retrospective observational study of patients diagnosed with COVID-19 and admitted from April 17, 2020 to November 19, 2020 in the RDT General Hospital in Bathalapalli, Anantapur District, Andhra Pradesh, India. During this time, the hospital was designated as a COVID-19 centre and was utilized exclusively to treat patients who had a positive SARS-Cov-2 reverse transcriptase polymerase chain reaction (RT-PCR) or antigen test. During this time, to reduce the risk of transmission in the community, patients with mild symptoms were also admitted for isolation. As per Government rules, even patients with mild symptoms could not be discharged at least until 10 days passed from the symptom onset or the first positive SARS-CoV-2 test.

For the study, we used routinely collected data (demographics, laboratory investigations, and vitals) entered in the hospital information system (HIS). Comorbidities of patients were not entered in the HIS and, therefore, could not be used in the prognostic models. The study was performed according to the principles of the Declaration of Helsinki. The associated Ethics Committee approved the study and waived the need for informed consent. The methodology of the study followed the guidelines for transparent reporting of a multivariable prediction model for individual prediction or diagnosis (TRIPOD) [5]. For the sample size, we took a practical approach by using all available data to maximize the power of the statistical analysis [6].

### Outcome and independent predictors

We aimed to develop a prognostic model with variables collected at the time of hospital admission that could be utilized to identify COVID-19 patients who were at higher risk of complications. We decided to use a composite endpoint including in-hospital mortality or hypoxaemia as the primary outcome of the study. Hypoxaemia was defined as having oxygen saturation below 93% or the need of oxygen support to maintain saturation above 93% [2, 7]. We selected *a priori* set of potential predictors according the availability of data in the HIS and whether the variables had shown to influence the outcome of COVID-19 in previous studies [8–13].

### Model development and validation

The dataset was split in two. Development of the model was performed with data from patients admitted from April 17 to August 31, 2020 (development cohort), while model validation was performed with patients admitted from September 1 to November 19, 2020 (validation cohort). Assuming missing at random, missing values were imputed using chained equations in 10 datasets, each with 10 iterations [14, 15]. The outcome was included as a predictor in the imputation of the development cohort but not in the validation cohort.

Because our primary objective was to develop a pragmatic bedside risk score that did not demand complex calculations, we decided to categorize continuous variables [12]. Model development was performed in four stages. In the first stage, we made an initial selection of predictors based on the goodness of fit between the outcome and predictors using generalised additive models (GAM).

Categorical predictors were entered as linear components in the models, and continuous predictors were smoothed using penalised thin plate splines [16]. In the second stage, we selected optimal cut-off values to categorize continuous variables based on visual inspection of the GAM models [17]. In the third stage, to reduce the risk of model overfitting, we used least absolute shrinkage and selection operator (LASSO) regression with theory-driven penalization to select predictors and their cut-off values to be included in the final model [18]. In the fourth stage, we used the coefficients from the logistic regression models to construct the prognostic index.

One common problem when comparing laboratory data is that laboratory values are highly dependent of the methodology used, and data normalization is needed. Usually, clinical laboratories have normal ranges that enclose 95% of values in a healthy population. In settings where the laboratory normal range differs substantially from our values, we recommend using the lower or upper normal limits of their laboratory as the reference to calculate the prognostic scores, although other forms of normalization are also possible [19].

Predicted probabilities of the outcome in the development and validation cohorts were calculated by fitting logistic regression models with the prognostic score as the only independent variable in each imputed dataset, and using Rubin’s rules to combine the results [15]. Discrimination of the prognostic index was assessed using the area under the receiver operating characteristic curve (AUROCC) with confidence intervals (CI) obtained through 2000 bootstrap samples [5]. Calibration was assessed with the Brier score and graphically by inspecting the smoothed relationship between the predicted and observed risk [5]. Clinical utility was assessed using decision curve analysis [20]. As the concept of net benefit can be difficult to grasp [21], we described an hypothetical scenario where the prognostic index was used to decide whether patients needed admission in the hospital. Risk groups were formed based on the predicted probability of the outcome: low risk (<5%), intermediate-low risk (5-10%), intermediate-high risk (10-20%), high risk (20-40%) and very high risk (>40%). We performed several sensitivity analyses. We checked the performance of the model using complete case data, segregated by gender, and using mortality as the outcome.

## Results

### Model development

During the study period, 6123 patients with COVID-19 were admitted in the hospital (Figure 1). Forty-two patients were excluded, 4035 patients were included in the development cohort and 2046 in the validation cohort. The overall average hospital length of stay was 6.92 days (median 6, interquartile range [IQR] 4 to 8). The median age was 48 years (IQR 34 to 59) and 2348 (38.6%) were female. Differences between the development and the validation cohort are described in Table 1. The primary outcome occurred in 961 (23.8%) patients in the development cohort and in 548 (26.8%) in the validation cohort.

**Figure 1.**
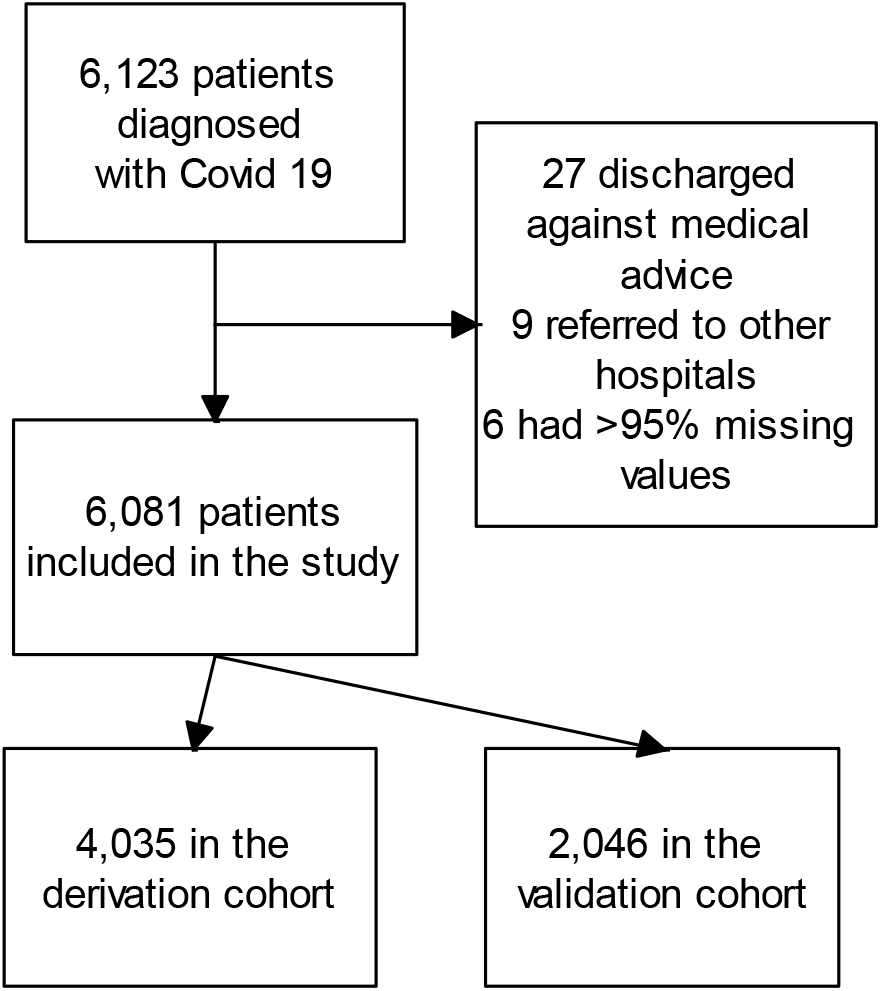
Flowchart of patients.

**Table 1.** Characteristics of patients in the development and the validation cohorts.

|  | Development cohort |  | Validation cohort |  |
| --- | --- | --- | --- | --- |
|  | Median (IQR)<br>or No (%) | Missing | Median (IQR)<br>or No (%) | Missing |
| Age-years | 48 (34-60) | 9 | 47 (33-58) | 0 |
| Systolic BP-mm Hg | 120 (110-120) | 1111 | 120 (110-120) | 4 |
| Diastolic BP-mm Hg | 80 (70-80) | 1111 | 80 (70-80) | 4 |
| Heart rate-min | 88 (82-92) | 1112 | 86 (80-90) | 5 |
| Respiratory rate-min | 20 (20-22) | 1117 | 20 (20-22) | 5 |
| Temperature-°F | 99 (99-99) | 1110 | 99 (98-99) | 5 |
| AST-IU/L | 25 (18-36) | 16 | 22 (17-31) | 3 |
| ALT-IU/L | 27 (18-42) | 18 | 26 (18-40) | 3 |
| Albumin-g/dL | 4.5 (4.2-4.8) | 18 | 4.6 (4.3-4.9) | 3 |
| LDH- IU/L | 385 (269-495) | 7 | 375 (284-467) | 1 |
| Creatinine-mg/dL | 0.8 (0.7-1) | 6 | 0.7 (0.6-0.8) | 3 |
| Urea-mg/dL | 22 (17-29) | 7 | 22 (18-29) | 3 |
| C-reactive protein-mg/dL | 0.5 (0.2-3) | 5 | 0.5 (0.3-2.8) | 1 |
| Sodium-mmol/L | 141 (139-143) | 5 | 141 (139-143) | 3 |
| Haemoglobin-g/dL | 13 (12-14) | 82 | 13 (12-14) | 0 |
| Platelet count-$\times 10^9/L$ | 265 (207-333) | 82 | 304 (243-364) | 0 |
| White cell count-$\times 10^9/L$ | 7 (5.5-9.1) | 82 | 6.9 (5.4-9) | 0 |
| Neutrophil count-$\times 10^9/L$ | 4.5 (3.3-6.2) | 82 | 4.4 (3.3-6.2) | 0 |
| Lymphocyte count-$\times 10^9/L$ | 1.9 (1.4-2.6) | 82 | 1.9 (1.4-2.4) | 0 |
| Neutrophil/Lymphocyte ratio | 2.3 (1.6-3.7) | 82 | 2.3 (1.6-3.8) | 0 |
| Female gender | 1593 (39.5) | 0 | 752 (36.8) | 0 |
| Deaths | 172 (4.3) | 0 | 53 (2.6) | 0 |
| Hypoxaemia | 959 (23.8) | 0 | 545 (26.6) | 0 |

IQR, interquartile range; BP, blood pressure; ALT, Alanine transaminase; AST, Aspartate transaminase; LDH, Lactate dehydrogenase

The model development is described in detail in the Supplementary Material. From the initial 20 predictor candidates, seven were excluded at the initial stage (Table S1). All remaining predictors were continuous variables, and were categorized using GAM to select the optimal cut-off values (Figures S1-S3 and Table S2). The final selection of cut-off values and variables was performed using LASSO logistic regression (Table S3), and coefficients were used to produce the prognostic scores (Table S4). The prognostic index ranged from 0 to 32 and included 12 variables: age, systolic blood pressure, heart rate, respiratory rate, aspartate transaminase, lactate dehydrogenase, urea, C-reactive protein, sodium, absolute lymphocyte count, absolute neutrophil count and neutrophil/lymphocyte ratio (Table 2). Table 2 also includes the reference range in our laboratory and suggested normalization values based on the upper and lower normal limits.

**Table 2.** The RCOS prognostic index.

|  | Reference range | Prognostic score |
| --- | --- | --- |
| **Age (years)** |  |  |
| 40-49 |  | 1 |
| 50-59 |  | 2 |
| 60-69 |  | 3 |
| >=70 |  | 4 |
| **Systolic BP (mm Hg) >= 140** |  | 1 |
| **Heart rate (pm) >=100** |  | 1 |
| **Respiratory rate (pm) >=22** |  | 2 |
| **AST-IU/L** | 0 to 40 |  |
| 40-79 | 1 to 2 x UNL | 1 |
| >=80 | >2 x UNL | 2 |
| **LDH- IU/L** | 207 to 414 |  |
| 700-899 | 1.69 to 2.17 x UNL | 1 |
| >=900 | >2.17 x UNL | 2 |
| **Urea-mg/dL** | 15 to 39 |  |
| 40-49.9 | 1 to 1.25 x UNL | 2 |
| >=50 | >1.25 x UNL | 3 |
| **C-reactive protein-mg/dL** | 0 to 0.5 |  |
| 0.5-0.9 | 1 to 1.99 x UNL | 1 |
| 1-1.9 | 2 to 3.99 x UNL | 2 |
| 2-3.9 | 4 to 7.99 x UNL | 3 |
| 4-5.9 | 8 to 11.99 x UNL | 4 |
| 6-8.9 | 12 to 17.99 x UNL | 5 |
| 9-11.9 | 18 to 23.99 x UNL | 6 |
| >=12 | >=24 x UNL | 7 |
| **Sodium-mmol/L <135** | 135 to 148 | 1 |
| **Lymphocyte count-$\times 10^9/L$** | 1 to 5 |  |
| <0.8 |  | 3 |
| 0.8-0.999 |  | 1 |
| **Neutrophil count-$\times 10^9/L$** | 1.2 to 8 |  |
| 8 - 9.9 |  | 1 |
| >=10 |  | 2 |
| **Neutrophil/Lymphocyte ratio** |  |  |
| 3-3.9 |  | 1 |
| 4-5.9 |  | 2 |
| 6-7.9 |  | 3 |
| >=8 |  | 4 |

BP, blood pressure; pm, per minute; AST, Aspartate transaminase; LDH, Lactate dehydrogenase; UNL, upper normal limit; LNL lower normal limit.

In the development cohort, the AUROCC was 0.907 (95% CI, 0.896-0.918) (Figure S4) and the Brier score was 0.0935. Calibration-in-the-large was 0.01 and the slope was 1.007 (Figure S5). Based on the predicted probability of the outcome we created the following risk groups: low risk (index 0, 1, or 2), intermediate-low risk (index 3 or 4), intermediate-high risk (index 5 or 6), high risk (index 7 or 8) and very high risk (index 9 or above) (Table 3).

**Table 3.**
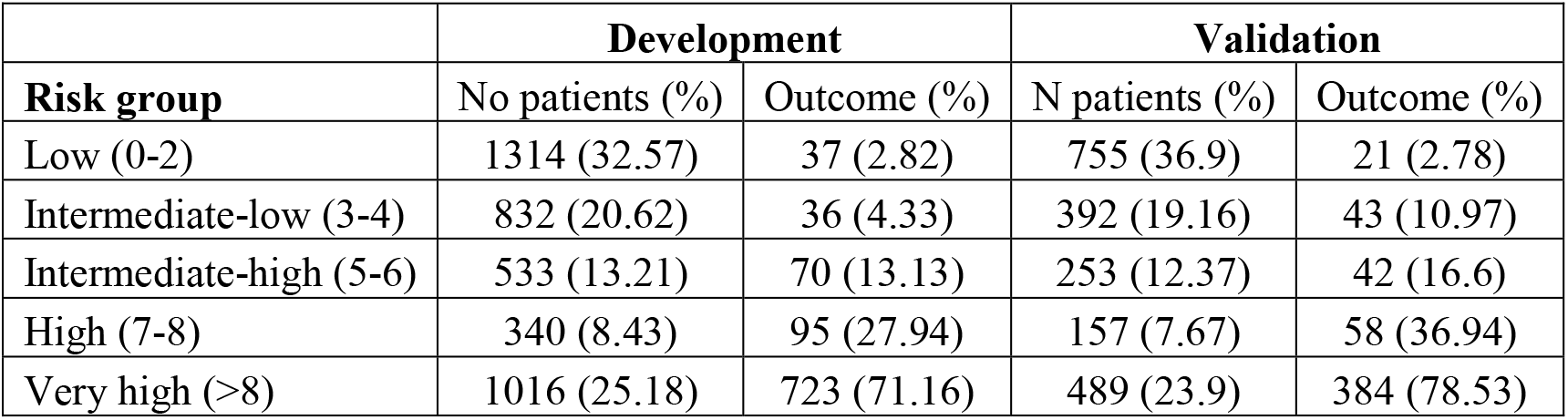
Proportion of patients who experienced the study outcome (death or hypoxaemia) segregated by risk group.

| **Risk group** | **Development** |  | **Validation** |  |
| --- | --- | --- | --- | --- |
|  | No patients (%) | Outcome (%) | N patients (%) | Outcome (%) |
| Low (0-2) | 1314 (32.57) | 37 (2.82) | 755 (36.9) | 21 (2.78) |
| Intermediate-low (3-4) | 832 (20.62) | 36 (4.33) | 392 (19.16) | 43 (10.97) |
| Intermediate-high (5-6) | 533 (13.21) | 70 (13.13) | 253 (12.37) | 42 (16.6) |
| High (7-8) | 340 (8.43) | 95 (27.94) | 157 (7.67) | 58 (36.94) |
| Very high (>8) | 1016 (25.18) | 723 (71.16) | 489 (23.9) | 384 (78.53) |

### Model validation

In the validation cohort, the AUROCC was 0.907 (95% CI, 0.892-0.922) and the Brier score was 0.098 (Figure 2 upper panel). Calibration-in-the-large was 0 (95% CI, −0.14 to 0.14) and the slope was 1 (95% CI, 0.91-1.09) (Figure 2 middle panel). Nearly 50% of cases had a prognostic index of 3 or less (Figure 2 lower panel and Table S5). The predicted risk for the primary outcome increased rapidly for prognostic scores between 5 and 10, and then had a progressive reduction (Figure 2 lower panel, Figure S6 and Table S5). Sensitivity, specificity, negative predictive value and positive predictive value of the prognostic model in the validation cohort are presented in Figure 3. In general, the proportion of patients with outcome by risk group was slightly larger than in the development cohort (Table 3).

**Figure 2.**
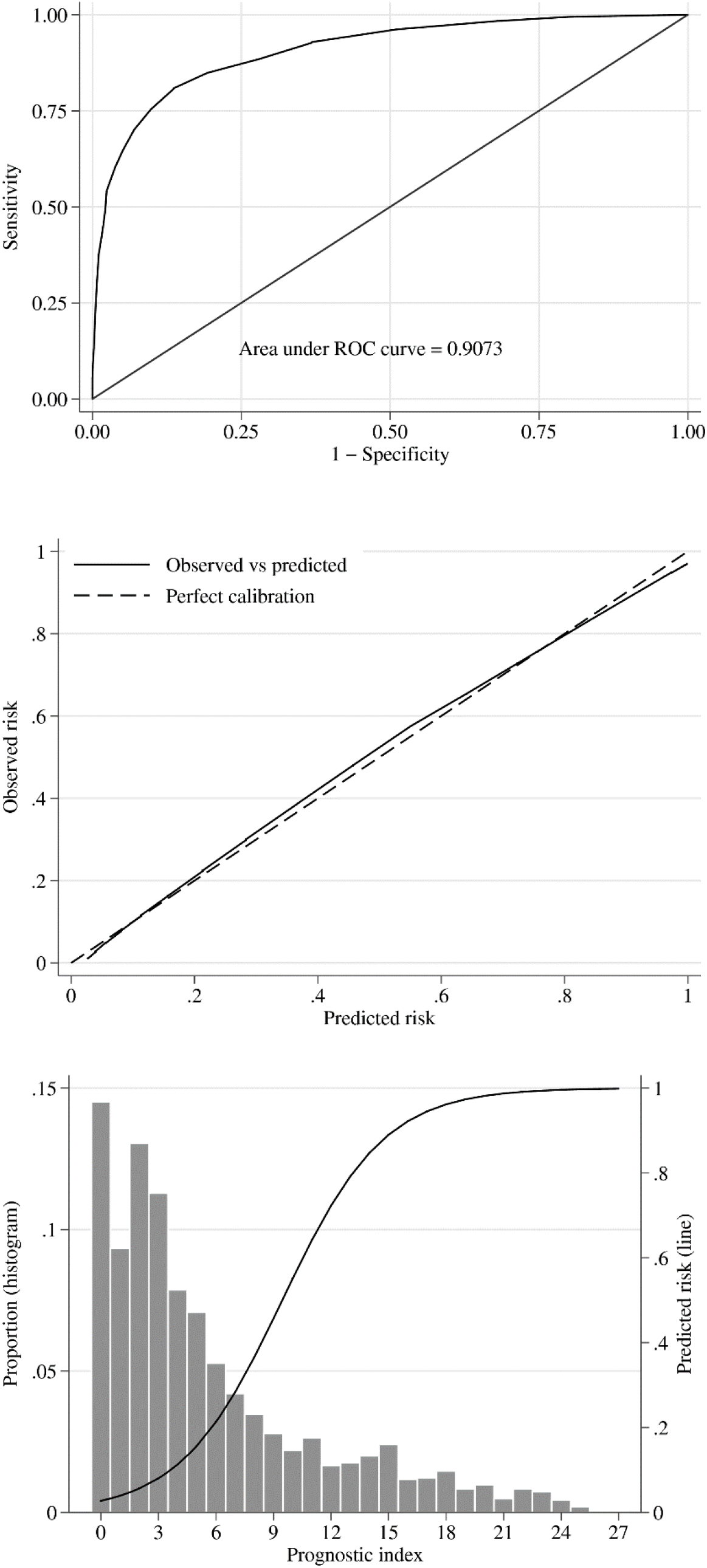
Discrimination (upper panel), calibration (middle panel), distribution of cases (lower panel histogram) and predicted probability of death or hypoxaemia (lower panel line) of the predictive model in the validation cohort.

**Figure 3.**
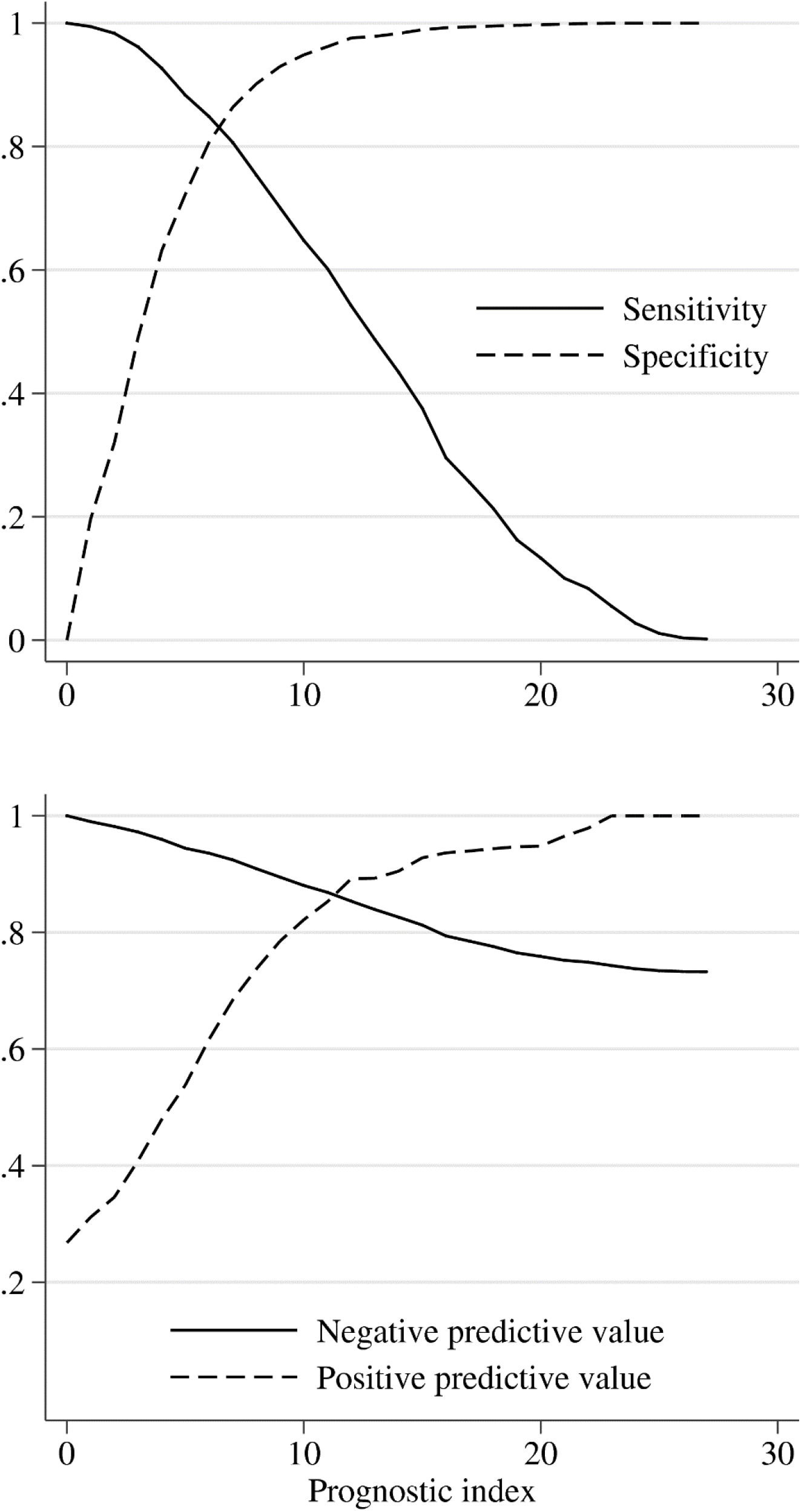
Sensitivity, specificity, negative predictive value and positive predictive value in the validation cohort.

Decision curve analysis is reported in Figure 4. The net benefit describes the performance of the model to identify true positives over true negatives [21]. Decisions curve analysis can be used to decide whether to initiate an intervention (e.g. to start a particular medication or to request a diagnostic test). However, to better understand the clinical use of the predictive model, we created a hypothetical scenario where patients were admitted (the “intervention”) according to the predicted probability of the outcome given by the prognostic index. We compared the performance of the model with two other possible scenarios: admit all patients (unlimited resources) or admit none (there are no free beds in the hospital). Actually, these two scenarios represent extreme situations that can occur in the real world. If the bed occupancy is high because of a sudden spike in the number of COVID-19 cases, we could select a higher prognostic index threshold for admission to optimize resources. If the incidence of COVID-19 cases comes down and the bed occupancy is low, we could be more permissible and reduce the prognostic score cut-off for admission. The selection of the threshold probability represents the trade-off between the benefit and the cost of the intervention. The net benefit of the predictive model was positive and above the net benefit of other alternatives (admit all or admit none) up to threshold probabilities above 90%, which are hardly justifiable in real life (Figure 4 upper panel). If we consider patients who did not develop the outcome did not need admission, the numbers of unnecessary admissions avoided is presented in the lower panel of Figure 4. For example, using a prognostic index of 7 or more as the threshold for admission, we could have reduced nearly 50% the number of the admissions.

**Figure 4.**
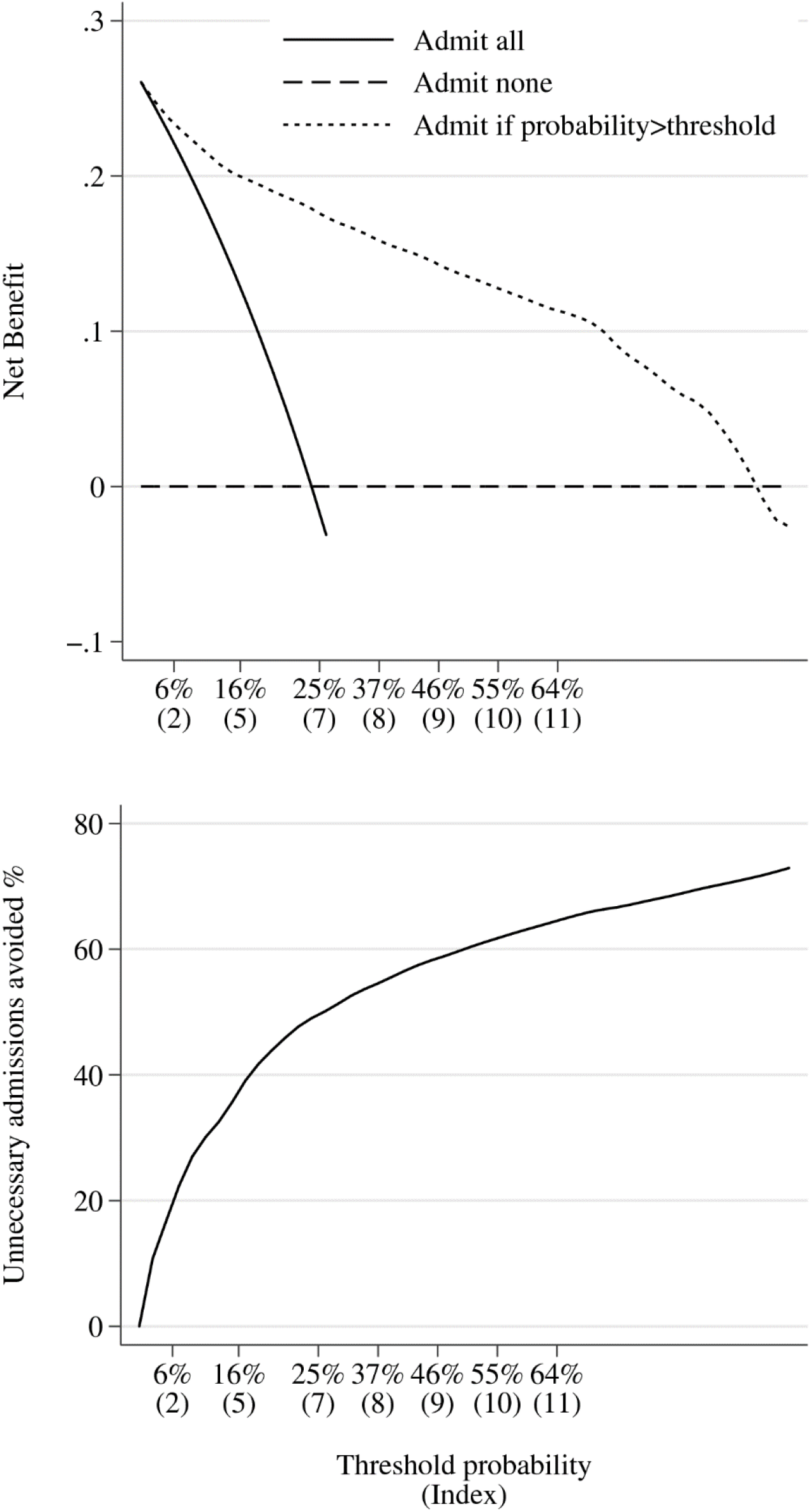
Decision curves. Net benefit (upper panel) and number of intervention avoided (lower panel) in the validation cohort.

In a sensitivity analysis using only complete cases, the results were almost identical (AUROCC was 0.907, 95% CI 0.893-0.922); Brier score 0.0977). The model performed slightly better in female cases (AUROCC 0.921, 95% CI, 0.896-0.945; Brier score 0.078) than in male cases (AUROCC 0.899, 95% CI 0.88-0.918; Brier score 0.109). The prognostic index showed excellent accuracy (AUROCC 0.947; 95% CI, 0.925-0.97) and calibration (Brier score 0.0188; calibration-in-the-large 0.0006, 95% CI, −0.315 to 0.316; slope 1.001, 95% CI, 0.816-1.188) to predict mortality. The performance of the model to predict mortality is described graphically in Figure 5 and Figures S7-S8.

**Figure 5.**
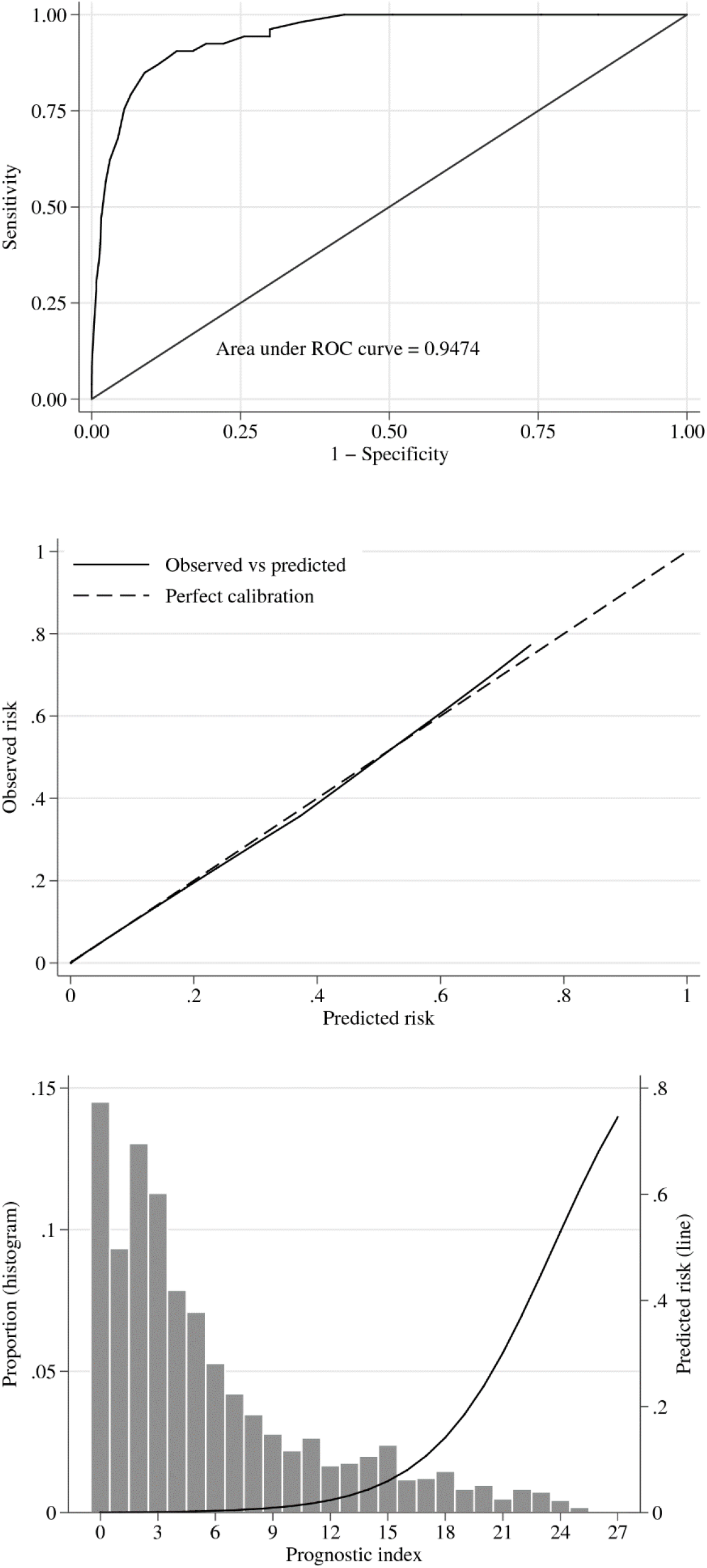
Discrimination (upper panel), calibration (middle panel), distribution of cases (lower panel histogram) and predicted probability (lower panel line) of the model to predict mortality in the validation cohort.

## Discussion

In this study, we present a pragmatic multivariable prognostic index that showed good discrimination calibration, and clinical utility in a cohort that could be considered representative of COVID-19 cases diagnosed in the community. The variables included in the RCOS prognostic index are readily available in most healthcare settings with basic laboratory infrastructure, and the index does not require complex calculation. It could be used to optimize resources in overwhelmed health systems, identifying patients who are more likely to develop complications and need hospital admission or closer ambulatory monitoring. Optimal utilization of resources is especially important in low and middle income countries, where public health facilities are overburden and unable to accommodate the high number of cases who need hospitalisation in the current COVID-19 pandemic.

Current therapy of COVID-19 focus on patients who have already developed complications [22]. Previous studies have shown that the highest level of viral replication occurs around the first day of symptoms, and 95% of hospitalized patients have negative viral cultures after 15 days of symptoms [23]. Current evidence suggests that antiviral and antibody therapy are more effective if started early, during the first days of symptoms [24, 25]. The RCOS prognostic index could be used to escalate therapy in patients with higher risk of complications. The index could also help identify high risk groups in targeted randomized clinical trials investigating early interventions aimed to reduce morbidity or mortality of COVID-19.

### Strengths and limitations

In COVID-19 cases, hypoxemia usually appears within five to ten days of symptoms [26–28]. In our study, patients were admitted regardless of the severity of symptoms and were not discharged before ten days passed from symptom onset. In the development cohort, 23.8% of the patients developed hypoxaemia or died, which is similar to the proportion of severe cases found in a large cohort from China [2]. This suggests that the model was developed in a population representative of COVID-19 in the community. However, the validation cohort had a larger proportion of patients with hypoxaemia than the development cohort, and both predicted and observed risk were higher than expected in some of the risk groups, suggesting a selection of more severe cases in the validation cohort as the clinical pressure to be admitted increased because the number of COVID-19 cases spiked in the region during the study period. When implementing the prognostic index in populations with lower (e.g. primary health centre) or higher (e.g. emergency department) expected risk, the use of risk groups may overestimate (primary health centre) or underestimate (emergency department) the real risk of complications. Although classifying patients in risk groups can still be useful as an initial reference, our results suggest that users of the prognostic index should try to estimate the predicted probability of the outcome in their settings.

The prognostic model was not developed to predict complications in hospital settings with high mortality. Still, the excellent performance of the prognostic index to predict mortality suggests that it could be a helpful companion to other severity predictors such as oxygen saturation or PaO2/FiO2 ratio to identify patients who are more likely to require ventilator or critical care support [29], but new studies are needed to confirm this hypothesis.

The study has several limitations. Unlike other COVID-19 prognostic models, the RCOS prognostic index does not include comorbid conditions of the patients [4]. It is possible that including comorbidity predictors could improve the performance of the model. This a single centre study, and validation was performed in the same setting as the development. However, we used data from a different period of time to validate our model, which is a stronger approach compared to other forms of internal validation, and can be considered intermediate between internal and external validation [5].

## Conclusion

Prognostic models are able to transform complex clinical situations into a single dimension numerical value. In this study, we present a prognostic score that demonstrated excellent discrimination and calibration to predict complications and mortality in a population of COVID-19 cases that included a large proportion of mild cases. If our results are validated in other settings, the RCOS prognostic index could help optimize resources in overstretched healthcare systems and improve clinical decisions in COVID-19 patients diagnosed in the community who are at higher risk of developing complications.

## Supporting information

TRIPOD

## Data Availability

The conditions of the ethical approval for the study preclude open access data sharing to reduce the risk of patient identification. Specific requests for data sharing will be considered subject to ethical approval and data transfer agreements.

## References

1. Wiersinga WJ, Rhodes A, Cheng AC, Peacock SJ, Prescott HC. Pathophysiology, Transmission, Diagnosis, and Treatment of Coronavirus Disease 2019 (COVID-19): A Review. JAMA 2020; 324: 782–793.

2. Wu Z, McGoogan JM. Characteristics of and Important Lessons From the Coronavirus Disease 2019 (COVID-19) Outbreak in China: Summary of a Report of 72?314 Cases From the Chinese Center for Disease Control and Prevention. JAMA 2020; 323: 1239–1242.

3. Collins GS, Reitsma JB, Altman DG, Moons KGM. Transparent Reporting of a multivariable prediction model for Individual Prognosis or Diagnosis (TRIPOD): the TRIPOD statement. Ann. Intern. Med. 2015; 162: 55–63.

4. Wynants L, Van Calster B, Collins GS, Riley RD, Heinze G, Schuit E, Bonten MMJ, Dahly DL, Damen JAA, Debray TPA, de Jong VMT, De Vos M, Dhiman P, Haller MC, Harhay MO, Henckaerts L, Heus P, Kammer M, Kreuzberger N, Lohmann A, Luijken K, Ma J, Martin GP, McLernon DJ, Andaur CL, Reitsma JB, Sergeant JC, Shi C, Skoetz N, Smits LJM, et al. Prediction models for diagnosis and prognosis of covid-19 infection: systematic review and critical appraisal. BMJ 2020; 369: m1328.

5. Moons KGM, Altman DG, Reitsma JB, Ioannidis JPA, Macaskill P, Steyerberg EW, Vickers AJ, Ransohoff DF, Collins GS. Transparent Reporting of a multivariable prediction model for Individual Prognosis or Diagnosis (TRIPOD): explanation and elaboration. Ann. Intern. Med. 2015; 162: W1–73.

6. Riley RD, Ensor J, Snell KIE, Harrell FE, Martin GP, Reitsma JB, Moons KGM, Collins G, van Smeden M. Calculating the sample size required for developing a clinical prediction model. BMJ 2020; 368: m441.

7. Lee WW, Mayberry K, Crapo R, Jensen RL. The accuracy of pulse oximetry in the emergency department. Am. J. Emerg. Med. 2000; 18: 427–431.

8. Cai Q, Huang D, Yu H, Zhu Z, Xia Z, Su Y, Li Z, Zhou G, Gou J, Qu J, Sun Y, Liu Y, He Q, Chen J, Liu L, Xu L. COVID-19: Abnormal liver function tests. J. Hepatol. 2020; 73: 566–574.

9. King JT, Yoon JS, Rentsch CT, Tate JP, Park LS, Kidwai-Khan F, Skanderson M, Hauser RG, Jacobson DA, Erdos J, Cho K, Ramoni R, Gagnon DR, Justice AC. Development and validation of a 30-day mortality index based on pre-existing medical administrative data from 13,323 COVID-19 patients: The Veterans Health Administration COVID-19 (VACO) Index. PloS One 2020; 15: e0241825.

10. Liang W, Liang H, Ou L, Chen B, Chen A, Li C, Li Y, Guan W, Sang L, Lu J, Xu Y, Chen G, Guo H, Guo J, Chen Z, Zhao Y, Li S, Zhang N, Zhong N, He J, China Medical Treatment Expert Group for COVID-19. Development and Validation of a Clinical Risk Score to Predict the Occurrence of Critical Illness in Hospitalized Patients With COVID-19. JAMA Intern. Med. 2020; 180: 1081–1089.

11. Satici C, Demirkol MA, Sargin Altunok E, Gursoy B, Alkan M, Kamat S, Demirok B, Surmeli CD, Calik M, Cavus Z, Esatoglu SN. Performance of pneumonia severity index and CURB-65 in predicting 30-day mortality in patients with COVID-19. Int. J. Infect. Dis. IJID Off. Publ. Int. Soc. Infect. Dis. 2020; 98: 84–89.

12. Knight SR, Ho A, Pius R, Buchan I, Carson G, Drake TM, Dunning J, Fairfield CJ, Gamble C, Green CA, Gupta R, Halpin S, Hardwick HE, Holden KA, Horby PW, Jackson C, Mclean KA, Merson L, Nguyen-Van-Tam JS, Norman L, Noursadeghi M, Olliaro PL, Pritchard MG, Russell CD, Shaw CA, Sheikh A, Solomon T, Sudlow C, Swann OV, Turtle LC, et al. Risk stratification of patients admitted to hospital with covid-19 using the ISARIC WHO Clinical Characterisation Protocol: development and validation of the 4C Mortality Score. BMJ 2020; 370: m3339.

13. Gupta RK, Harrison EM, Ho A, Docherty AB, Knight SR, van Smeden M, Abubakar I, Lipman M, Quartagno M, Pius R, Buchan I, Carson G, Drake TM, Dunning J, Fairfield CJ, Gamble C, Green CA, Halpin S, Hardwick HE, Holden KA, Horby PW, Jackson C, Mclean KA, Merson L, Nguyen-Van-Tam JS, Norman L, Olliaro PL, Pritchard MG, Russell CD, Scott-Brown J, et al. Development and validation of the ISARIC 4C Deterioration model for adults hospitalised with COVID-19: a prospective cohort study. Lancet Respir. Med. 2021;.

14. Wood AM, White IR, Royston P. How should variable selection be performed with multiply imputed data? Stat. Med. 2008; 27: 3227–3246.

15. Morris TP, White IR, Royston P. Tuning multiple imputation by predictive mean matching and local residual draws. BMC Med. Res. Methodol. 2014; 14: 75.

16. Wood SN. Generalized additive models: an introduction with R. CRC press; 2017.

17. Barrio I, Arostegui I, Quintana JM, Group I-C. Use of generalised additive models to categorise continuous variables in clinical prediction. BMC Med. Res. Methodol. 2013; 13: 83.

18. Ahrens A, Hansen CB, Schaffer ME. lassopack: Model selection and prediction with regularized regression in Stata. Stata J. SAGE Publications Sage CA: Los Angeles, CA; 2020; 20: 176–235.

19. Karvanen J. The Statistical Basis of Laboratory Data Normalization. Drug Inf. J. SAGE Publications; 2003; 37: 101–107.

20. Vickers AJ, Elkin EB. Decision curve analysis: a novel method for evaluating prediction models. Med. Decis. Mak. Int. J. Soc. Med. Decis. Mak. 2006; 26: 565–574.

21. Vickers AJ, van Calster B, Steyerberg EW. A simple, step-by-step guide to interpreting decision curve analysis. Diagn. Progn. Res. 2019; 3: 18.

22. Therapeutic Management of Adults With COVID-19 [Internet]. COVID-19 Treat. Guidel. [cited 2021 Mar 19].Available from: https://www.covid19treatmentguidelines.nih.gov/therapeutic-management/.

23. van Kampen JJA, van de Vijver DAMC, Fraaij PLA, Haagmans BL, Lamers MM, Okba N, van den Akker JPC, Endeman H, Gommers DAMPJ, Cornelissen JJ, Hoek RAS, van der Eerden MM, Hesselink DA, Metselaar HJ, Verbon A, de Steenwinkel JEM, Aron GI, van Gorp ECM, van Boheemen S, Voermans JC, Boucher CAB, Molenkamp R, Koopmans MPG, Geurtsvankessel C, van der Eijk AA. Duration and key determinants of infectious virus shedding in hospitalized patients with coronavirus disease-2019 (COVID-19). Nat. Commun. 2021; 12: 267.

24. Cohen MS. Monoclonal Antibodies to Disrupt Progression of Early Covid-19 Infection. N. Engl. J. Med. 2021; 384: 289–291.

25. Beigel JH, Tomashek KM, Dodd LE, Mehta AK, Zingman BS, Kalil AC, Hohmann E, Chu HY, Luetkemeyer A, Kline S, Lopez de Castilla D, Finberg RW, Dierberg K, Tapson V, Hsieh L, Patterson TF, Paredes R, Sweeney DA, Short WR, Touloumi G, Lye DC, Ohmagari N, Oh M-D, Ruiz-Palacios GM, Benfield T, Fätkenheuer G, Kortepeter MG, Atmar RL, Creech CB, Lundgren J, et al. Remdesivir for the Treatment of Covid-19 - Final Report. N. Engl. J. Med. 2020; 383: 1813–1826.

26. Wang D, Hu B, Hu C, Zhu F, Liu X, Zhang J, Wang B, Xiang H, Cheng Z, Xiong Y, Zhao Y, Li Y, Wang X, Peng Z. Clinical Characteristics of 138 Hospitalized Patients With 2019 Novel Coronavirus-Infected Pneumonia in Wuhan, China. JAMA 2020; 323: 1061–1069.

27. Huang C, Wang Y, Li X, Ren L, Zhao J, Hu Y, Zhang L, Fan G, Xu J, Gu X, Cheng Z, Yu T, Xia J, Wei Y, Wu W, Xie X, Yin W, Li H, Liu M, Xiao Y, Gao H, Guo L, Xie J, Wang G, Jiang R, Gao Z, Jin Q, Wang J, Cao B. Clinical features of patients infected with 2019 novel coronavirus in Wuhan, China. Lancet Lond. Engl. 2020; 395: 497–506.

28. Zhou F, Yu T, Du R, Fan G, Liu Y, Liu Z, Xiang J, Wang Y, Song B, Gu X, Guan L, Wei Y, Li H, Wu X, Xu J, Tu S, Zhang Y, Chen H, Cao B. Clinical course and risk factors for mortality of adult inpatients with COVID-19 in Wuhan, China: a retrospective cohort study. Lancet Lond. Engl. 2020; 395: 1054–1062.

29. Gupta RK, Marks M, Samuels THA, Luintel A, Rampling T, Chowdhury H, Quartagno M, Nair A, Lipman M, Abubakar I, van Smeden M, Wong WK, Williams B, Noursadeghi M, UCLH COVID-19 Reporting Group. Systematic evaluation and external validation of 22 prognostic models among hospitalised adults with COVID-19: an observational cohort study. Eur. Respir. J. 2020; 56.

