## Supplementary Material for "Development and validation of the RCOS prognostic index: a bedside multivariable logistic regression model to predict hypoxaemia or death in patients with SARS-CoV-2 infection"

### Software

Statistical analysis was performed using Stata 14.2, except for generalised additive models (GAM),which were performed in R (version 4.0.3) with the mgcv library (version 1.8-33) [1]. Missing data were handled by multiple imputation using chained equations and predictive mean matching (with five nearest neighbours) for continuous variables in 10 datasets, each with 10 iterations [2, 3]. The selection of variables for the final model was performed using least absolute shrinkage and selection operator (LASSO) logistic regression with theory-driven penalization, which have shown to reduce the risk of overfitting compared with other penalization methods (rlassologit command from the lassopack version 1.4.1) [4]. Calibration slopes were calculated with the coefficient of a logistic model for the outcome and the model linear predictor as the independent variable; and calibration-in-the-large was calculated with the intercept of a logistic model for the outcome with the model linear predictor as an offset term.

### Model development

Our aim was to construct a simple predictive score that could be used bedside by clinicians without the need of computers or mobile applications. The model was developed in four stages.

#### Stage 1

We assessed the goodness of fit between the outcome and predictors using GAM models. We excluded predictors with a deviance explained below 2% (female gender, diastolic blood pressure, temperature, haemoglobin concentration and platelet count). To avoid multicollinearity problems, when two predictors were highly correlated (such as Alanine transaminase (ALT) and Aspartate transaminase (AST); and white cell count and neutrophil count), we excluded the ones that had lower goodness of fit (ALT and white cell count).

**Table S1.**

| Predictors | $R^2$ | DE | REML | |
|---|---|---|---|---|
| Female gender | 0.011 | 1.1% | 2195.1 | Excluded |
| Age-years | 0.107 | 10.7% | 1979.4 | |
| Systolic BP-mm Hg | 0.050 | 4.0% | 1754.8 | |
| Diastolic BP-mm Hg | 0.009 | 0.8% | 1810.2 | Excluded |
| Heart rate-min | 0.030 | 2.4% | 1781.4 | |
| Respiratory rate-min | 0.103 | 8.0% | 1675.4 | |
| Temperature-°F | 0.017 | 1.6% | 1802.0 | Excluded |
| AST-IU/L | 0.084 | 7.3% | 2057.6 | |
| ALT-IU/L | 0.041 | 3.7% | 2136.7 | Excluded |
| Albumin-g/dL | 0.144 | 13.1% | 1925.6 | |
| LDH- IU/L | 0.147 | 12.1% | 1959.7 | |
| Creatinine-mg/dL | 0.076 | 6.8% | 2072.6 | |
| Urea-mg/dL | 0.169 | 13.8% | 1919.9 | |
| C-reactive protein-mg/dL | 0.370 | 32.0% | 1518.9 | |
| Sodium-mmol/l | 0.156 | 13.4% | 1933.0 | |
| Haemoglobin-g/dL | 0.002 | 0.3% | 2185.6 | Excluded |
| Platelet count-×10^9/L | 0.010 | 0.9% | 2174.8 | Excluded |
| White cell count-×10^9/L | 0.081 | 6.4% | 2056.5 | Excluded |
| Neutrophil count-×10^9/L | 0.129 | 10.3% | 1970.5 | |
| Lymphocyte count-×10^9/L | 0.165 | 14.2% | 1886.9 | |
| Neutrophil/Lymphocyte ratio | 0.272 | 23.1% | 1691.2 | |

DE, deviance explained, REML, restricted maximum likelihood; BP, blood pressure; ALT, Alanine transaminase; AST, Aspartate transaminase; LDH, Lactate dehydrogenase.

#### Stage 2

We selected optimal cut-off values to categorize continuous variables based on visual inspection of the GAM models [5], taking into account clinically important points, the laboratory reference range for normal values (we avoided cut-off values within the normal range), cut-off values used in other risk scores, and the distribution of values in the dataset (we avoided placing cut-off values far below the percentile 5 or far above the percentile 95). To minimise the loss of information produced by categorizing continuous variables, we tried to keep similar “risk-distance” between cut-off values (Figures S1-S3 and Table S2).

**Figure S1.** GAM models 1. DE=deviance explained. Vertical lines represent percentile 50 (solid line); 25 and 75 (dashed line ----); 10 and 90 (dotted line …..); 5 and 95 (dashed/dotted lines _._._). Risk denotes the log-odds of the outcome (hypoxaemia or death).

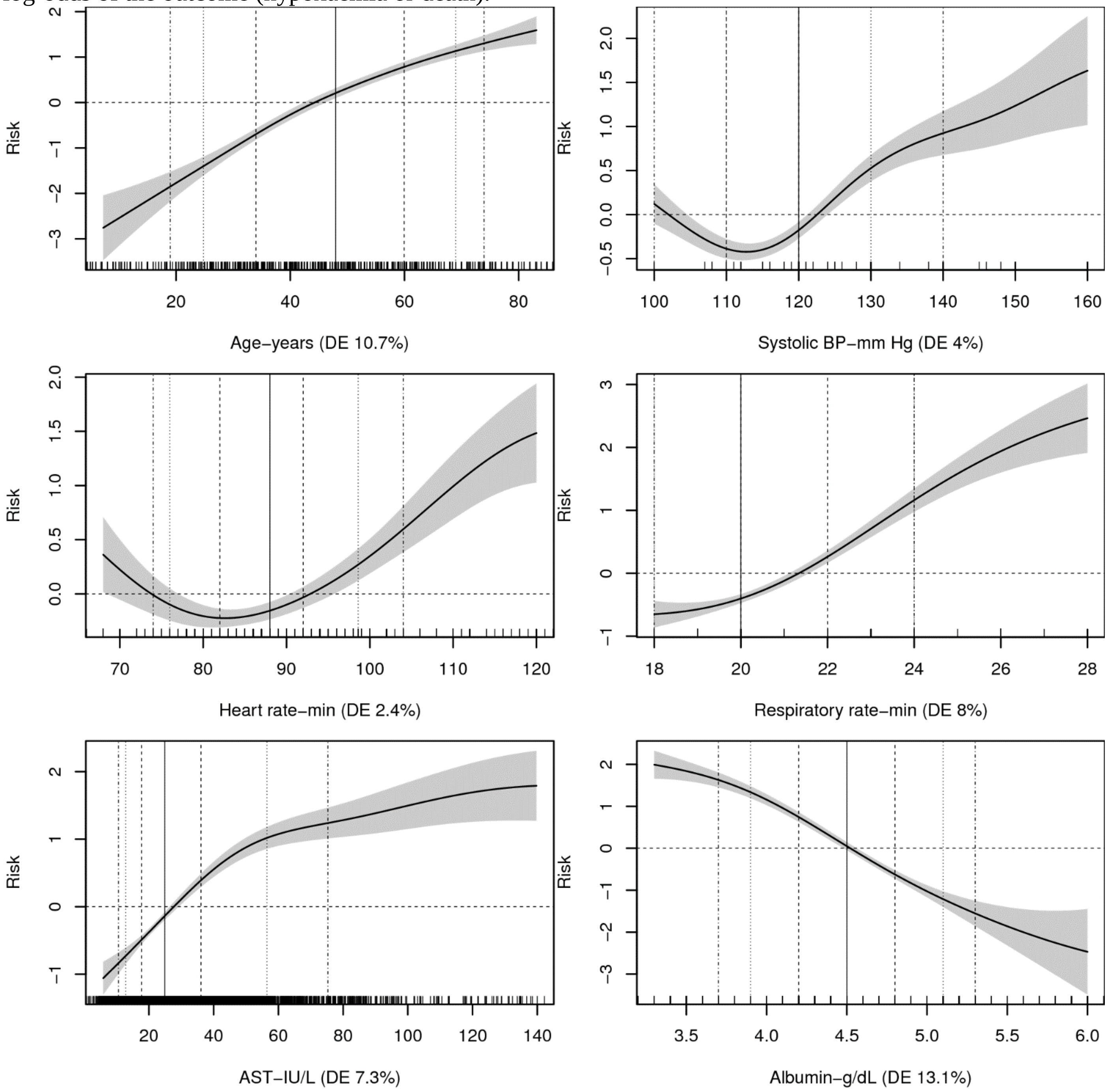

**Figure S2.** GAM models 2. DE=deviance explained. Vertical lines represent percentile 50 (solid line); 25 and 75 (dashed line ----); 10 and 90 (dotted line …..); 5 and 95 (dashed/dotted lines _._._)

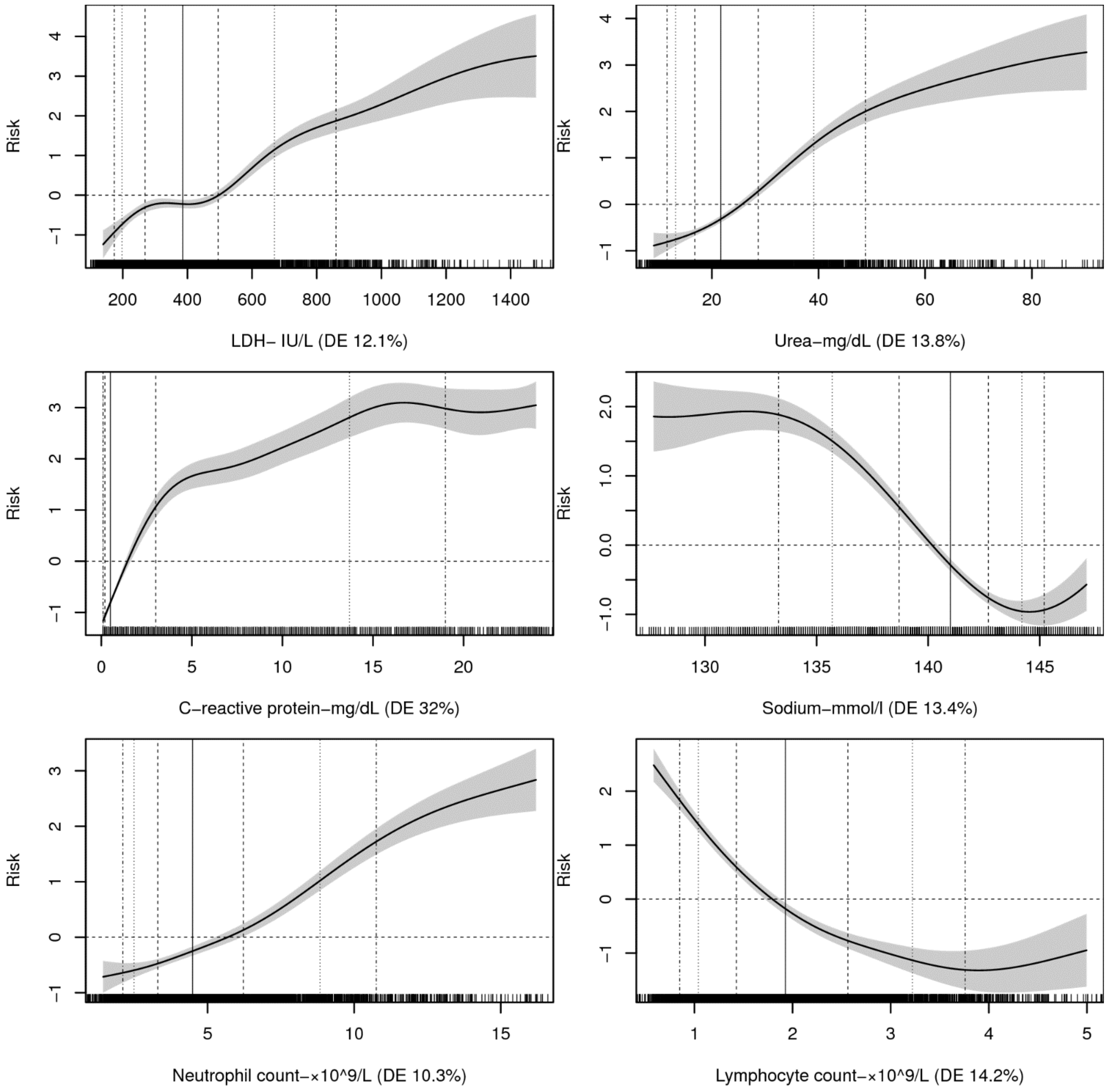

**Figure S3**. GAM models 3. DE=deviance explained. Vertical lines represent percentile 50 (solid line); 25 and 75 (dashed line ----); 10 and 90 (dotted line …..); 5 and 95 (dashed/dotted lines _._._)

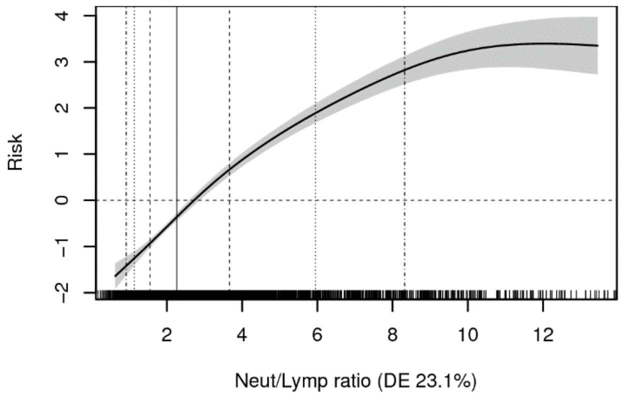

**Table S2.** Selected cut-off values for continuous variables.

| | Laboratory NR | Cut-off |
|---|---|---|
| Age-years | NA | 40,50,60,70 |
| Systolic BP-mm Hg | NA | 140 |
| Heart rate-min | NA | 100 |
| Respiratory rate-min | NA | 22 |
| AST-IU/L | 0-40 | 40, 80 |
| Albumin-g/dL | 3.5-5.3 | 3.5 |
| LDH- IU/L | 207-414 | 400, 700, 900 |
| Urea-mg/dL | 15-39 | 40, 50 |
| C-reactive protein-mg/dL | 0-0.5 | 0.5,1,2,4,6,9,12 |
| Sodium-mmol/L | 135-148 | 135 |
| Neutrophil count-×10^9/L | 1.2-8 | 8, 10 |
| Lymphocyte count-×10^9/L | 1-5 | 0.8, 1 |
| Neutrophil/Lymphocyte ratio | NA | 3,4,6,8 |

NR, normal range; BP, blood pressure; AST, Aspartate transaminase; LDH, Lactate dehydrogenase

#### Stage 3

Using the initial cut-off values selected in Stage 2, we performed LASSO logistic regression for each imputed dataset looking for overall agreement to select the cut-off values for the final model [2]. There was a 100% agreement among the imputed datasets. All initial cut-off values were included by the LASSO model except albumin (which was excluded from the final model) and LDH 400 IU/L. See table S3.

**Table S3.** Selection of cut-off values using LASSO regression.

| | Initial selection (Stage 2) | LASSO selection |
|---|---|---|
| Age-years | 40,50,60,70 | 40,50,60,70 |
| Systolic BP-mm Hg | 140 | 140 |
| Heart rate-min | 100 | 100 |
| Respiratory rate-min | 22 | 22 |
| AST-IU/L | 40, 80 | 40, 80 |
| Albumin-g/dL | 3.5 | Excluded |
| LDH- IU/L | 400, 700, 900 | 700, 900 |
| Urea-mg/dL | 40, 50 | 40, 50 |
| C-reactive protein-mg/dL | 0.5,1,2,4,6,9,12 | 0.5,1,2,4,6,9,12 |
| Sodium-mmol/L | 135 | 135 |
| Neutrophil count-×10^9/L | 8, 10 | 8, 10 |
| Lymphocyte count-×10^9/L | 0.8, 1 | 0.8, 1 |
| Neutrophil/Lymphocyte ratio | 3,4,6,8 | 3,4,6,8 |

BP, blood pressure; AST, Aspartate transaminase; LDH, Lactate dehydrogenase

#### Stage 4

We combined logistic regression models from the imputed datasets using Rubin's rules. Coefficients from this logistic model and LASSO penalised coefficients were combined and scaled (x3) to produce the prognostic index.

**Table S4.** LASSO regression, logistic regression coefficients and final prognostic index.

| | Penalised coefficient | Logit coefficient (95% CI) | Prognostic score |
|---|---|---|---|
| Age (years) | | | |
| 40-49 | 0.16 | 0.65 (0.32 to 0.98) | 1 |
| 50-59 | 0.53 | 0.98 (0.67 to 1.29) | 2 |
| 60-69 | 0.76 | 1.24 (0.91 to 1.58) | 3 |
| >=70 | 0.87 | 1.38 (0.97 to 1.8) | 4 |
| Systolic BP (mm Hg) | | | |
| >= 140 | 0.32 | 0.39 (0.02 to 0.76) | 1 |
| Heart rate (pm) | | | |
| >=100 | 0.19 | 0.31 (-0.04 to 0.65) | 1 |
| Respiratory rate (pm) | | | |
| >=22 | 0.64 | 0.77 (0.54 to 1) | 2 |
| AST-IU/L | | | |
| 40-79 | 0.38 | 0.43 (0.17 to 0.69) | 1 |
| >=80 | 0.61 | 0.85 (0.38 to 1.32) | 2 |
| LDH- IU/L | | | |
| 700-899 | 0.3 | 0.37 (-0.05 to 0.8) | 1 |
| >=900 | 0.58 | 0.71 (0.2 to 1.23) | 2 |
| Urea-mg/dL | | | |
| 40-49.9 | 0.54 | 0.61 (0.21 to 1.01) | 2 |
| >=50 | 0.95 | 1.01 (0.55 to 1.48) | 3 |
| C-reactive protein-mg/dL | | | |
| 0.5-0.9 | 0.23 | 0.73 (0.37 to 1.09) | 1 |
| 1-1.9 | 0.61 | 1.04 (0.69 to 1.4) | 2 |
| 2-3.9 | 0.93 | 1.32 (0.96 to 1.68) | 3 |
| 4-5.9 | 1.44 | 1.83 (1.4 to 2.25) | 4 |
| 6-8.9 | 1.82 | 2.24 (1.79 to 2.69) | 5 |
| 9-11.9 | 2.05 | 2.47 (1.99 to 2.95) | 6 |
| >=12 | 2.46 | 2.8 (2.44 to 3.16) | 7 |
| Sodium-mmol/L | | | |
| <135 | 0.49 | 0.46 (0.14 to 0.79) | 1 |
| Lymphocyte count-×10^9/L | | | |
| <0.8 | 0.86 | 1.06 (0.54 to 1.58) | 3 |
| 0.8-0.99 | 0.24 | 0.32 (-0.11 to 0.75) | 1 |
| Neutrophil count-×10^9/L | | | |
| 8 - 9.9 | 0.1 | 0.25 (-0.17 to 0.66) | 1 |
| >=10 | 0.67 | 0.88 (0.37 to 1.39) | 2 |
| Neutrophil/Lymphocyte ratio | | | |
| 3-3.9 | 0.25 | 0.36 (0.07 to 0.65) | 1 |
| 4-5.9 | 0.47 | 0.52 (0.19 to 0.84) | 2 |
| 6-7.9 | 0.77 | 0.83 (0.32 to 1.35) | 3 |
| >=8 | 1.07 | 1.15 (0.52 to 1.77) | 4 |

### Model performance in the development cohort

**Figure S4.** Discrimination in the development cohort.

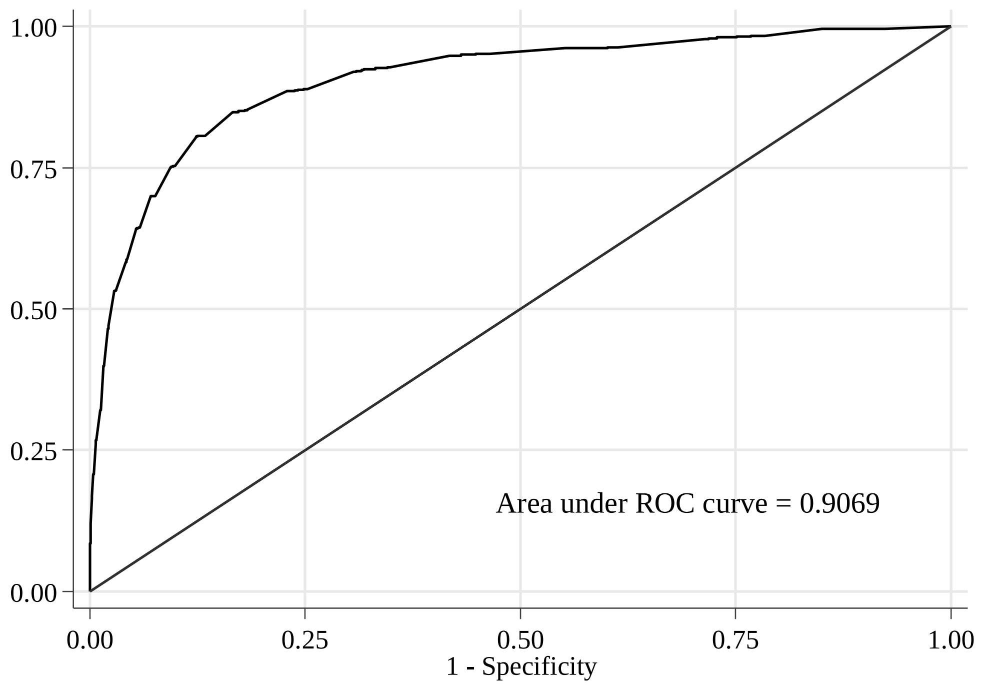

**Figure S5.** Calibration in the development cohort.

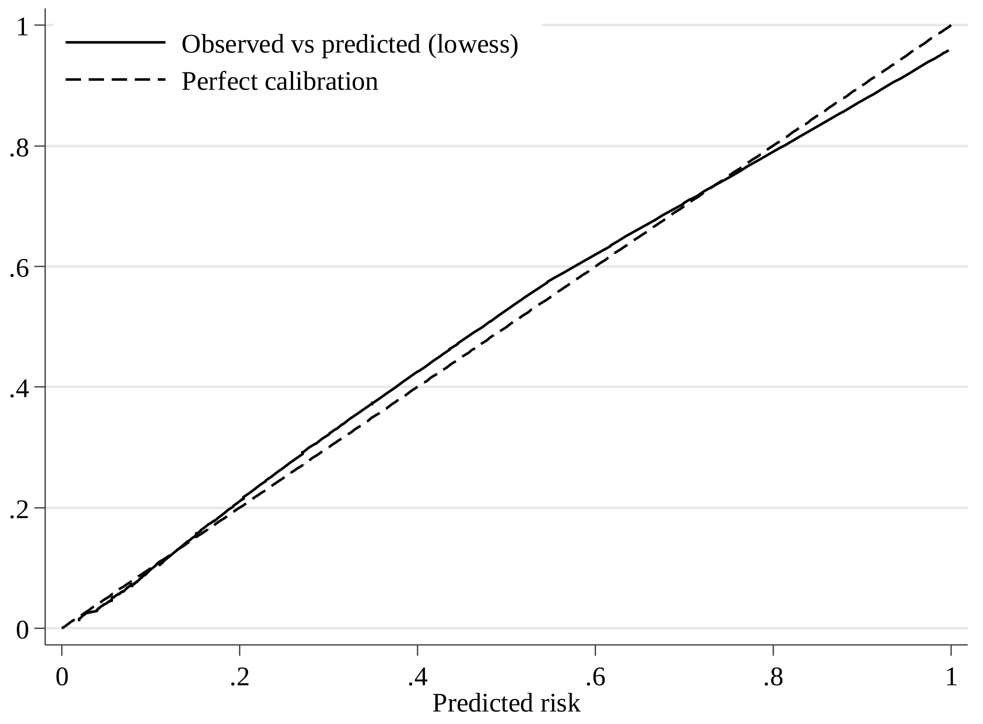

### Predicted risk given by prognostic scores in the validation cohort

**Table S5.** Distribution of patients and predicted risk of the outcome (hypoxaemia or death) and mortality in the validation cohort.

| | Distribution | | | Predicted risk | |
|---|---|---|---|---|---|
| Prognostic index | N | Percent | Cumulative percent | Outcome | Mortality |
| 0 | 297 | 14.52 | 14.52 | 2.74% | 0.05% |
| 1 | 191 | 9.34 | 23.85 | 3.95% | 0.07% |
| 2 | 267 | 13.05 | 36.9 | 5.66% | 0.10% |
| 3 | 231 | 11.29 | 48.19 | 8.05% | 0.14% |
| 4 | 161 | 7.87 | 56.06 | 11.31% | 0.19% |
| 5 | 145 | 7.09 | 63.15 | 15.68% | 0.26% |
| 6 | 108 | 5.28 | 68.43 | 21.35% | 0.35% |
| 7 | 86 | 4.2 | 72.63 | 28.34% | 0.49% |
| 8 | 71 | 3.47 | 76.1 | 36.58% | 0.67% |
| 9 | 57 | 2.79 | 78.89 | 45.68% | 0.92% |
| 10 | 45 | 2.2 | 81.09 | 55.09% | 1.26% |
| 11 | 54 | 2.64 | 83.72 | 64.14% | 1.73% |
| 12 | 34 | 1.66 | 85.39 | 72.28% | 2.36% |
| 13 | 36 | 1.76 | 87.15 | 79.18% | 3.23% |
| 14 | 41 | 2 | 89.15 | 84.72% | 4.39% |
| 15 | 49 | 2.39 | 91.54 | 88.99% | 5.94% |
| 16 | 24 | 1.17 | 92.72 | 92.18% | 8.00% |
| 17 | 25 | 1.22 | 93.94 | 94.50% | 10.70% |
| 18 | 30 | 1.47 | 95.41 | 96.16% | 14.16% |
| 19 | 17 | 0.83 | 96.24 | 97.34% | 18.51% |
| 20 | 20 | 0.98 | 97.21 | 98.16% | 23.83% |
| 21 | 10 | 0.49 | 97.7 | 98.73% | 30.11% |
| 22 | 17 | 0.83 | 98.53 | 99.13% | 37.23% |
| 23 | 15 | 0.73 | 99.27 | 99.40% | 44.95% |
| 24 | 9 | 0.44 | 99.71 | 99.59% | 52.93% |
| 25 | 4 | 0.2 | 99.9 | 99.72% | 60.76% |
| 26 | 1 | 0.05 | 99.95 | 99.81% | 68.07% |
| 27 | 1 | 0.05 | 100 | 99.87% | 74.59% |

**Figure S6.** Percentage risk increase by one point increase in the prognostic index.

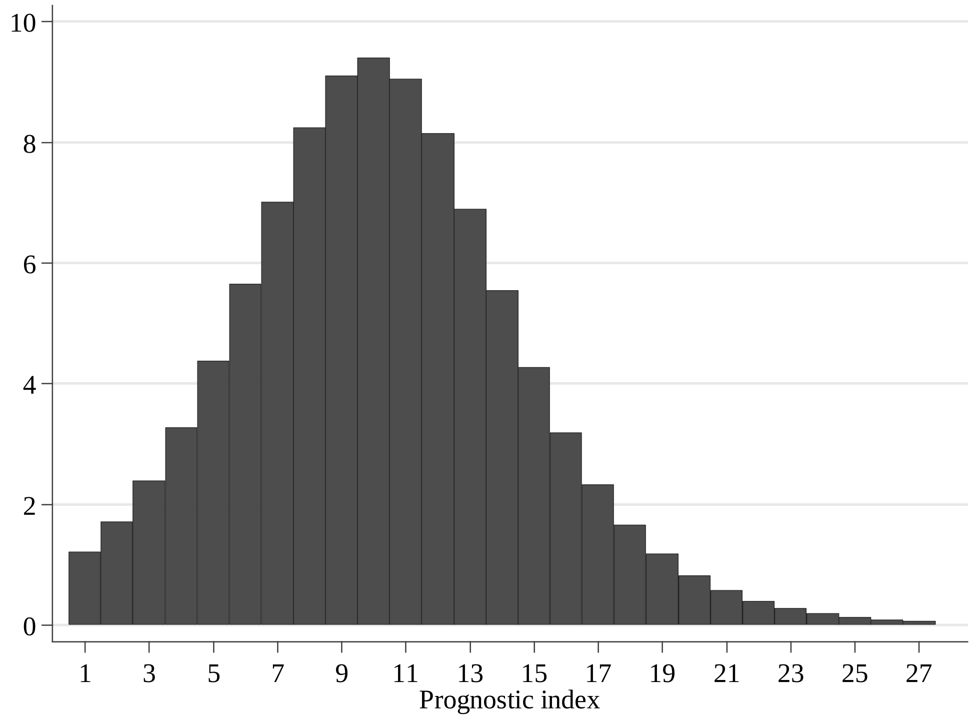

#### Model performance to predict mortality in the validation cohort

**Figure S7.** Sensitivity, specificity, negative predictive value and positive predictive value of the predictive model to predict mortality in the validation cohort.

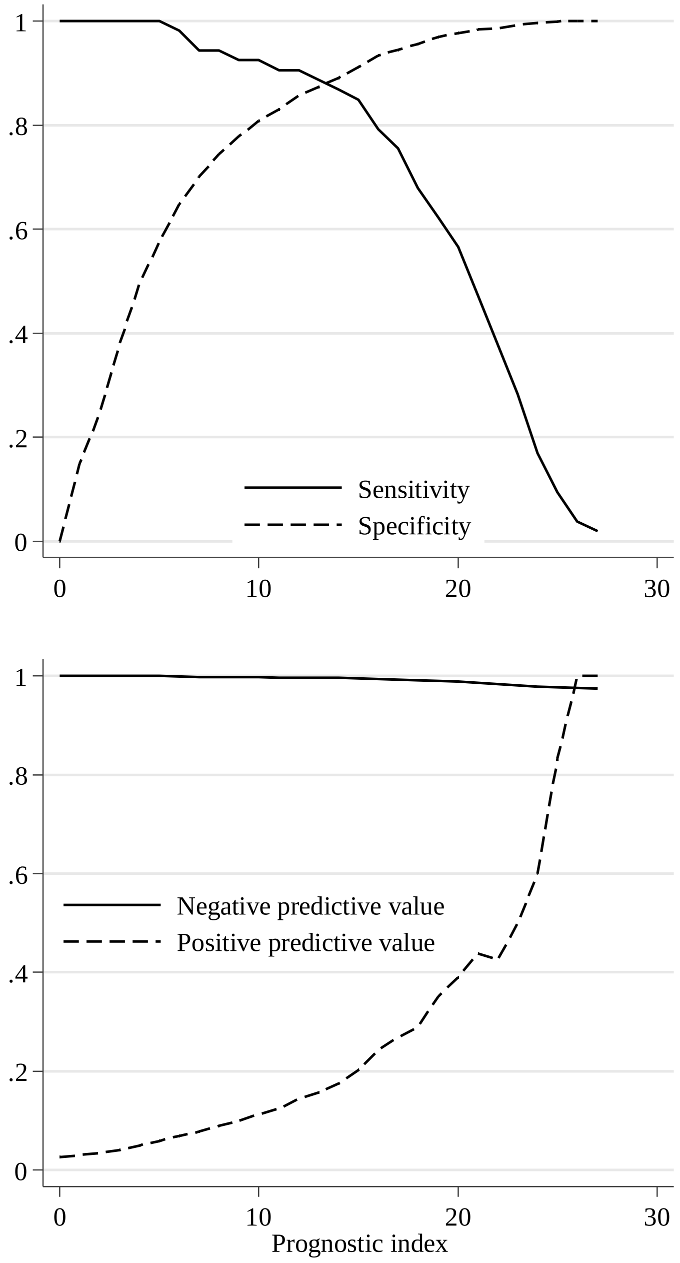

**Figure S8.** Decision curves. Net benefit (upper panel) and number of intervention avoided (lower panel) of the prognostic model for mortality in the validation cohort.

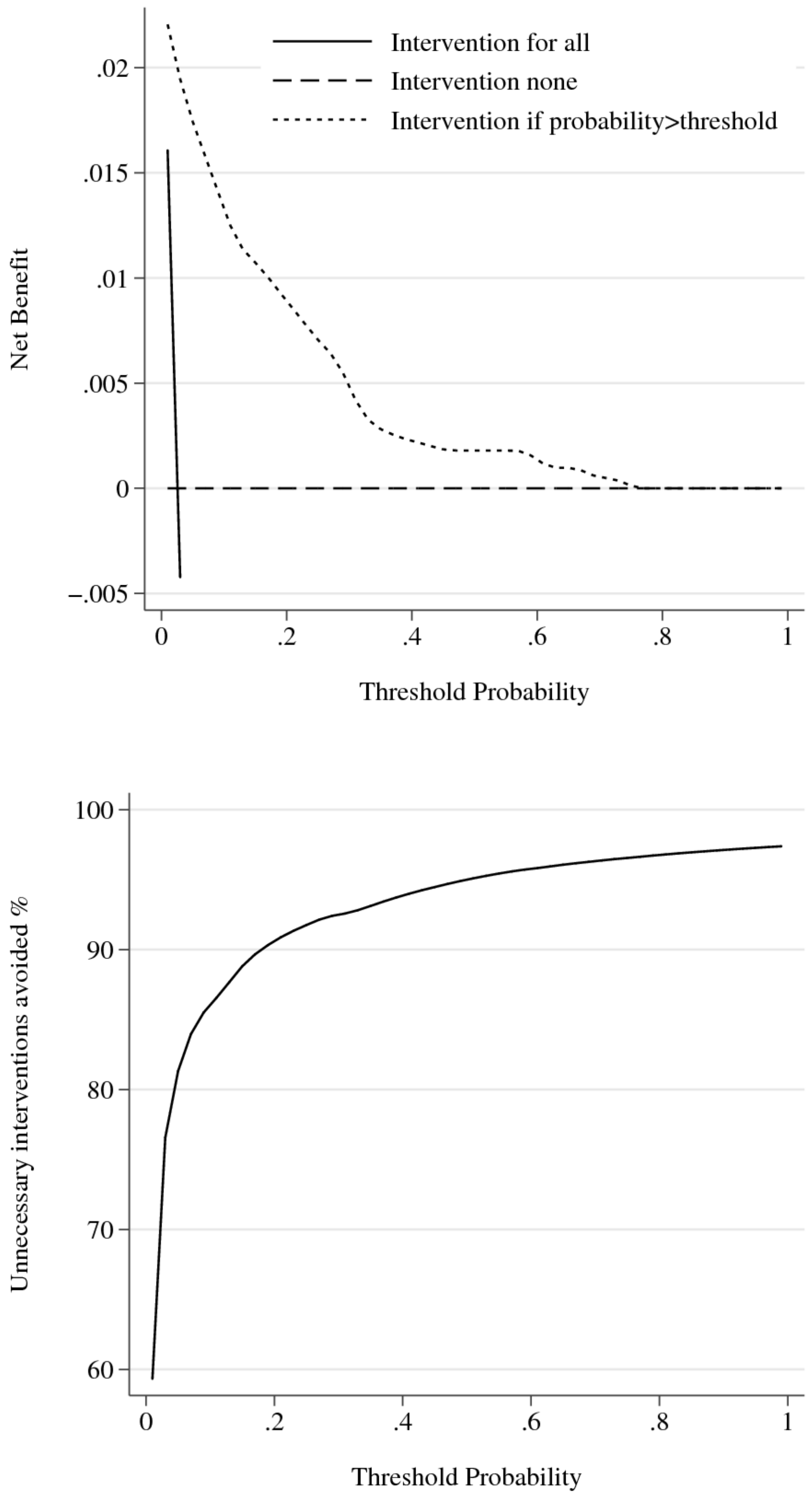

#### References

1. Wood SN. Generalized additive models: an introduction with R. CRC press; 2017.

2. Wood AM, White IR, Royston P. How should variable selection be performed with multiply imputed data? Stat Med. 2008;27:3227–46.

3. Morris TP, White IR, Royston P. Tuning multiple imputation by predictive mean matching and local residual draws. BMC Med Res Methodol. 2014;14:75.

4. Ahrens A, Hansen CB, Schaffer ME. lassopack: Model selection and prediction with regularized regression in Stata. Stata J. 2020;20:176–235.

5. Barrio I, Arostegui I, Quintana JM, Group I-C. Use of generalised additive models to categorise continuous variables in clinical prediction. BMC Med Res Methodol. 2013;13:83.
